# Crowd Annotations Can Approximate Clinical Autism Impressions from Short Home Videos with Privacy Protections

**DOI:** 10.1101/2021.07.01.21259683

**Authors:** Peter Washington, Emilie Leblanc, Kaitlyn Dunlap, Aaron Kline, Cezmi Mutlu, Brianna Chrisman, Nate Stockham, Kelley Paskov, Dennis P. Wall

## Abstract

Artificial Intelligence (A.I.) solutions are increasingly considered for telemedicine. For these methods to adapt to the field of behavioral pediatrics, serving children and their families in home settings, it will be crucial to ensure the privacy of the child and parent subjects in the videos. To address this challenge in A.I. for healthcare, we explore the potential for global image transformations to provide privacy while preserving behavioral annotation quality. Crowd workers have previously been shown to reliably annotate behavioral features in unstructured home videos, allowing machine learning classifiers to detect autism using the annotations as input. We evaluate this method with videos altered via pixelation, dense optical flow, and Gaussian blurring. On a balanced test set of 30 videos of children with autism and 30 neurotypical controls, we find that the visual privacy alterations do not drastically alter any individual behavioral annotation at the item level. The AUROC on the evaluation set was 90.0% +/- 7.5% for the unaltered condition, 85.0% +/- 9.0% for pixelation, 85.0% +/- 9.0% for optical flow, and 83.3% +/- 9.3% for blurring, demonstrating that an aggregation of small changes across multiple behavioral questions can collectively result in increased misdiagnosis rates. We also compare crowd answers against clinicians who provided the same annotations on the same videos and find that clinicians are more sensitive to autism-related symptoms. We also find that there is a linear correlation (r=0.75, p<0.0001) between the mean Clinical Global Impression (CGI) score provided by professional clinicians and the corresponding classifier score emitted by the logistic regression classifier with crowd inputs, indicating that the classifier’s output probability is a reliable estimate of clinical impression of autism from home videos. A significant correlation is maintained with privacy alterations, indicating that crowd annotations can approximate clinician-provided autism impression from home videos in a privacy-preserved manner.

## Introduction

Artificial intelligence (A.I.) is increasingly being considered to help healthcare solutions scale. In order for such solutions to be deployed in clinical settings, several issues beyond simply achieving high performance must be addressed. In particular, the pipelines which host A.I. models must be seamless and intuitive for all users, and the system must garner maximal trust from all stakeholders.

While A.I. approaches are needed for healthcare to achieve scale and consistency, current A.I.-powered solutions to diagnostics within psychiatry and behavioral sciences are under-performant due to the complexity of the underling behaviors. Until A.I. has advanced to a point where it can seamlessly pass the Turing Test [39] and understand human social behavior, with all of its subtleties and nuances, to a degree that surpasses an average human’s social acuity (i.e., to the level of a licensed clinician), it is unlikely that A.I. will replace any behavioral healthcare jobs.

In the meantime, A.I. solutions which can augment the capabilities of non-expert humans may allow for scalable and accessible remote diagnostics while easing the burden of professional clinicians and the healthcare system to provide in person initial consultations.

Privacy rises to the forefront of patient concern when humans are incorporated into the diagnostic pipeline [56]. Many parents are uncomfortable with sharing raw videos of their children with strangers online even if the purpose of sharing is to help the parent’s receive affordable and accessible diagnostic services [52]. However, human-in-the-loop techniques are pertinent given the current limitations of A.I., and privacy concerns must be directly addressed in any scalable solution where the humans in the loop are strangers (i.e., crowdsourced workers).

Here, we address these issues by studying an A.I.-augmented pipeline for detecting Autism Spectrum Disorder, or autism, from unstructured home videos. Autism is a developmental delay which is currently estimated to affect 1 in 40 children [27]. While access to care requires a formal diagnosis, access to diagnostic services is severely limited for many families, thus limiting potential care. Some evidence suggests that as much as 80% of families in the United States lack access to care [34], and underserved populations are disproportionately affected [17]. A.I.powered telemedical solutions therefore have the possibility to help these families, many of whom may otherwise lack access to traditional health services.

Crowd-powered telemedical diagnostic tools can provide parents with a risk score and an associated probability for a diagnosis, and prior works have repeatedly demonstrated the success of A.I. models to successfully detect autism using solely annotations provided by non-clinical human workers [5-8, 29, 37-38, 45, 47-48, 50-53, 57]. This family of solutions utilizes a distributed crowdsourced workforce to quickly annotate behavioral features displayed in videos recorded during parent-administered home autism therapy sessions using mobile digital health therapies [20-24] and wearable augmented reality solutions [2-4, 14-15, 26, 33, 40-43, 54-55].

In order for such solutions to truly scale for annotation by a large crowd workforce, the privacy of the patients must be preserved. This is important in general for healthcare applications, but it is especially critical when the patients in question are young children with a developmental delay and observed by a stranger in the privacy of their home. Our prior work has shown that applying light privacy-preserving modifications to videos, such as pitch shifting and covering the child’s face with a virtual box, results in minimal degradation of the quality of crowdsourced annotations used for remote detection of autism-related behaviors [52]. However, such lightweight privacy protections may be insufficient to some patients, such as those patients who do not want the interior of their home exposed to strangers on the Internet.

Here, we explore the effect of standard visual privacy-preserving mechanisms. In particular, we apply pixelation, dense optical flow, and Gaussian blurring, which are either standard methods for protecting the privacy of human subjects in image and video datasets [35] or for representing visual features for activity recognition [25] while obfuscating personally identifiable visual features. We study the effect of these video transformations on annotation quality. On a balanced test set of 30 videos of children with autism and 30 neurotypical controls, we find that no visual privacy condition deviates from the unaltered condition by more than half of a categorical ordinal severity point out of 4 questions corresponding to behavior severity. We compare crowd responses to professional clinicians and find that the probability score emitted by the classifier is consistent with the clinicians’ global impression from watching the same video. We also find that clinicians are more sensitive to autism-related symptoms than crowd workers.

## Methods

### Balanced video dataset

We leveraged a balanced video dataset of 30 children with autism and 30 controls without autism. Both groups were gender and aged matched: we posted 30 videos of male children (13 with autism and 17 neurotypical) and 30 videos of female children (17 with autism and 13 neurotypical). The mean age in the videos of children with autism was 3.49 years old (SD=1.58 years old), and the mean age in the videos of children without autism was 3.41 years old (SD=1.39 years old).

### Logistic regression classifier for predicting autism

We utilized a previously validated [37-38, 52] logistic regression classifier for predicting autism vs. not autism from the answers to the multiple-choice questions we asked crowd workers. The classifier [28, 30] was derived from the Autism Diagnostic Observation Schedule (ADOS) module 2 [31] and was trained on clinician filled scoresheets from the Boston Autism Consortium (AC), the Simons Simplex Collection v14 (SSC) [10], Autism Genetic Resource Exchange (AGRE) [11], National Database of Autism Research (NDAR) [16] and the Simons Variation in Individuals Project (SVIP) [36].

To generate confidence intervals, we performed 10,000 iterations for a bootstrapping procedure for each video. In each iteration, we sampled with replacement from the 60 videos used for evaluation and computed the accuracy, precision, recall / sensitivity, specificity, Area Under the Receiver Operating Characteristic (AUROC), and Area Under the Precision-Recall Curve (AUPRC) on the resulting video set.

### Privacy conditions

We evaluated three privacy modifications applied to each video: (1) pixelation, (2) dense optical flow, and (3) Gaussian blur. Pixelation and Gaussian blurring are common methods for protecting the privacy of human subjects in image and video datasets [35]. Dense optical flow is a standard method for representing visual features for activity recognition [25] which happens to also preserve the privacy of the human subject in the video. To apply pixelation, we first resized the input frames down to 32×32 pixels while applying bilinear interpolation. We then calculated the final pixelated frame by resizing the smaller frame back to the original frame size using bilinear interpolation. To calculate dense optical flow, we applied Farnebäck’s algorithm for two-frame motion estimation based on polynomial expansion [9]. The image was colored through obtaining a 2-channel array with optical flow vectors, where the direction of the vectors corresponds to the hue of the image and the magnitude of the vectors corresponds to the value (lightness) of the image. To apply Gaussian blurring, we used a blurring kernel with 0.25 the width and height of the input image.

### Crowd annotation

To recruit crowd workers, we posted a video rating task on the crowdsourcing website Microworkers.com [18]. In this video rating task, crowd workers are asked to answer a series of 13 multiple choice questions, with 4 answer choices each, about the child’s behavior exhibited in each of 8 video videos (4 featuring a child with autism and 4 featuring a child without autism).

We provided the categorical ordinal variables corresponding to each worker’s answers into a pretrained binary logistic regression classifier for autism (see subsection *Logistic regression classifier for predicting autism* below for details about classifier training). 1,000 crowd workers completed the task and passed basic quality control checks for answer acceptance. Quality control measures included time spent on the annotation task and deviations in answers between videos [52]. We did not include workers who did not pass these basic quality control checks in the list of 1,000 evaluated workers.

We measured the classifier’s prediction for all 8 filtering videos for all workers. We then measured the mean probability of the correct class (PCC) for each worker across all 8 videos, where the PCC is the classifier’s output probability *p* when the true class of a video is autism and *1-p* otherwise. Out of the 1,000 workers evaluated, exactly 40 workers had a mean PCC at or above 80% when the mean PCC was rounded up. We recruited these 40 crowd workers to rate the primary video set of 60 videos used in this study.

Each of the 40 crowd workers were tasked with rating all 60 videos described in the subsection *Balanced video dataset*. We randomly split the 40 workers into 4 groups, and each group was assigned to one privacy condition per video. Therefore, no worker saw more than one version of each video. Furthermore, all workers saw exactly 15 unaltered videos, 15 videos with pixelation, 15 videos with dense optical flow, and 15 videos with Gaussian blurring. This ensured that no privacy condition was biased by any crowd worker annotation biases.

### Clinician annotation

We recruited 19 clinicians to rate the same balanced video dataset and provide the same categorical ordinal video-wide annotations as the crowd workers. All clinicians were licensed professionals who provide diagnoses of autism as part of their job duties. All clinicians were asked an additional question which crowd workers were not asked: “*Do you think the child has autism?*” The answer choices were:

- No, I am confident the child does not have autism (0)
- No, but I am unsure (1)
- Yes, but I am unsure (2)
- Yes, I am confident the child has autism (3)

All 60 videos received at least 1 complete rating by at least 1 clinician. Some videos were rated by more than 1 clinician, in which case we recorded the mean of the clinician answers for that video. For the “*Do you think the child has autism?*” question, we coded the responses from 0 to 3 as shown above.

Clinicians were also asked to provide a Clinical Global Impression (CGI) [12] rating for the children in the videos. The CGI scale measures the “severity of illness” between 1 (“normal, not at all ill”) to 7 (“among the most extremely ill patients”) and is designed to allow clinicians to provide a global impression without providing a formal diagnosis.

### Item-level analysis

For each question we asked crowd workers, we measured the mean absolute deviation of the mean answer for each privacy condition from the mean answer for the baseline condition. This difference provides a measure of the privacy condition’s effect on annotation quality. We hypothesized that some questions would be more susceptible to alteration with certain privacy conditions applied.

## Results

All procedures performed in studies involving human participants were in accordance with the ethical standards of the institution (approved by the Stanford University Institutional Review Board) and with the 1964 Helsinki declaration and its later amendments or comparable ethical standards.

### Performance of clinicians and crowd workers

Out of the 30 videos of children with autism, clinicians rated 8 videos confidently (with a mean autism rating above 2.5 out of 3.0). By contrast, clinicians rated 18 of the videos of neurotypical children confidently (with a mean autism rating below 0.5 out of 3.0). All 8 confidently rated videos were of children with autism, while 16 of the 18 neurotypical children were correctly identified (only 2 were actually diagnosed with autism). This suggests that clinicians observing remote videos of children are cautious about calling an autism diagnosis, but when they do guess a diagnosis, the child is very likely to actually have autism.

The clinician’s classifier correctly identified 28 of the 30 autism cases while only correctly identifying 14 of the 30 neurotypical cases. By contrast, the crowd’s classifier correctly identified 25 of the 30 autism cases and 27 of the 30 neurotypical cases. This suggests that clinicians are more sensitive to autism-related symptoms than crowd workers, thus resulting in a higher frequency of autism diagnosis by the binary classifiers.

There is a clear linear correlation (r=0.75, p<0.001) between the mean Clinical Global Impression (CGI) score provided by professional clinicians for each video and the corresponding classifier score emitted by the logistic regression classifier with crowd inputs (Figure 1). This suggests that crowd responses in conjunction with machine learning algorithms can approximate clinician intuition, and the classifier’s output can be interpreted as a fairly reliable approximation of clinical global impressions of autism.

**Figure 1.**
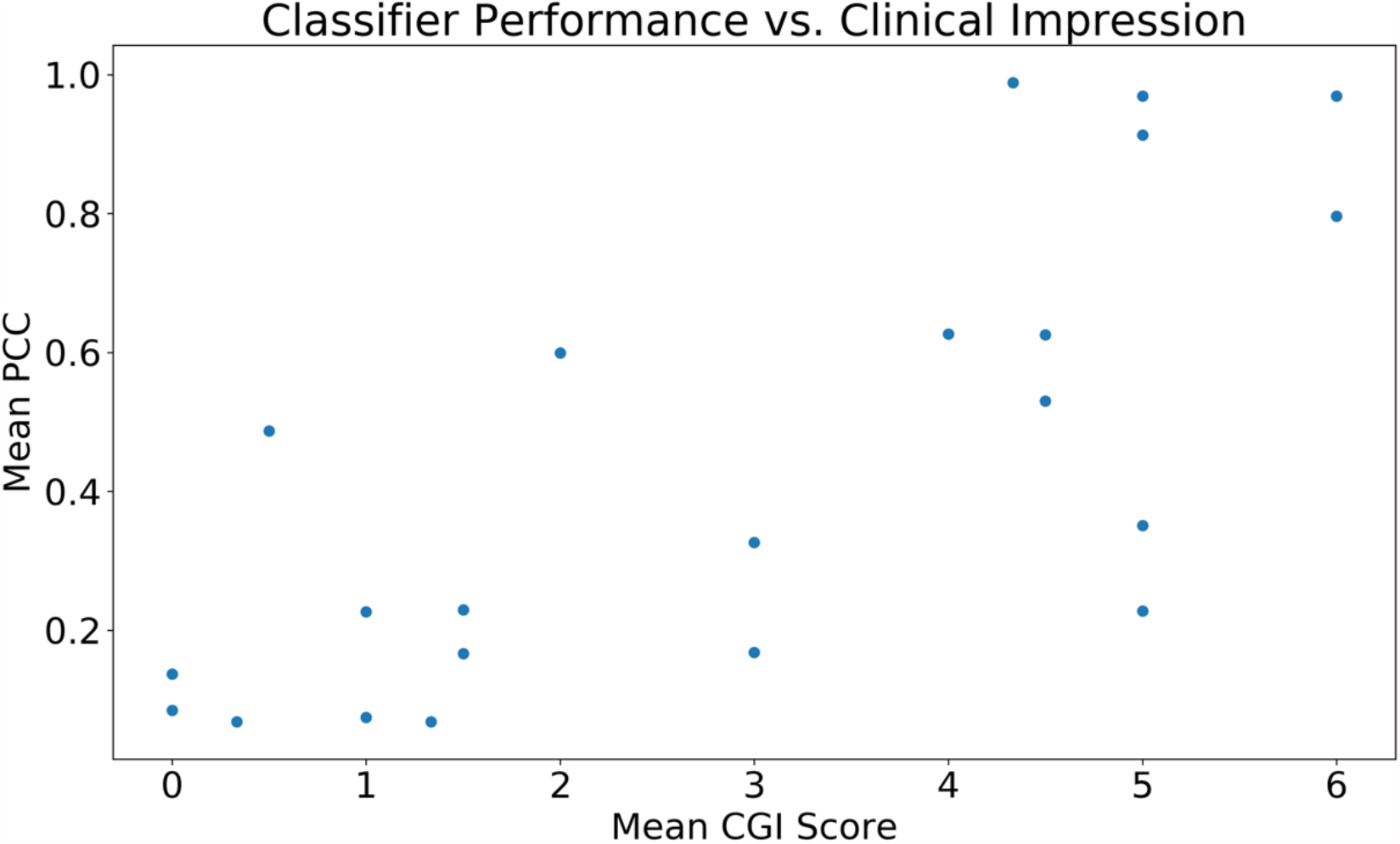
There is a clear linear correlation (r=0.75, p<0.001) between the mean Clinical Global Impression (CGI) score provided by professional clinicians for each video and the corresponding classifier score emitted by the logistic regression classifier with crowd inputs.

**Figure 2.**
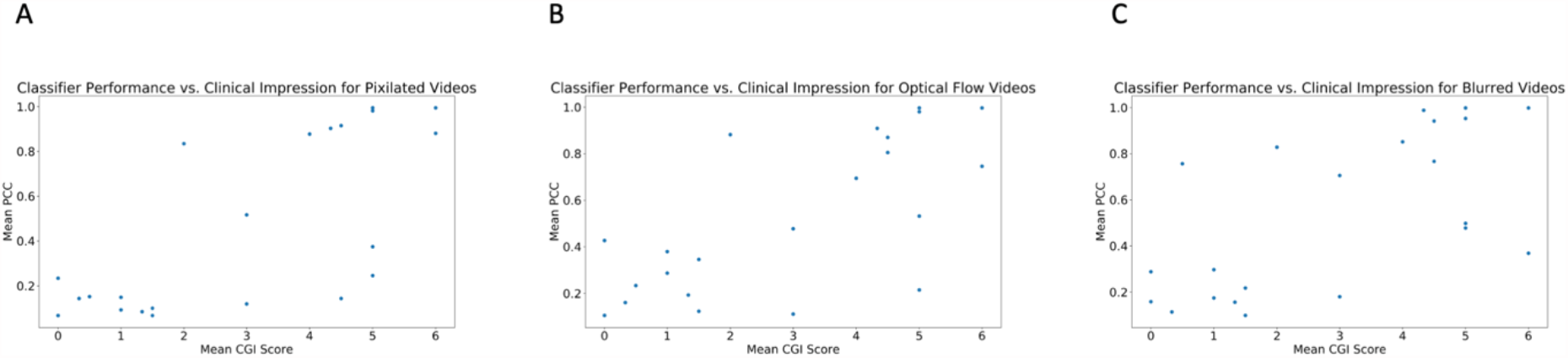
The linear correlation between the mean Clinical Global Impression (CGI) score provided by professional clinicians for each video and the corresponding classifier score emitted by the logistic regression classifier with crowd inputs is maintained with privacy-preserving video modifications. The correlation is weaker for Gaussian blurring (r=0.64, p=0.001 for Gaussian blurring) than for dense optical flow and pixelation (r=0.71, p=0.0002 for both).

Of the 60 unaltered videos, the mean performance metrics were 90.0% +/- 7.5% accuracy, 92.9% +/- 8.9% precision, 86.7% +/- 11.8% recall (sensitivity), and 93.3% +/- 8.6% specificity. The mean AUROC was 90.0% +/- 7.5% and the mean AUPRC was 93.1% +/- 6.2%.

### Effect of privacy conditions

The linear correlation between the mean Clinical Global Impression (CGI) score provided by professional clinicians for each video and the corresponding classifier score emitted by the logistic regression classifier with crowd inputs is maintained with privacy-preserving video modifications. The correlation is weaker for Gaussian blurring (r=0.64, p=0.001 for Gaussian blurring) than for dense optical flow and pixelation (r=0.71, p=0.0002 for pixelation for both).

With pixelation, the mean performance metrics were 85.0% +/- 9.2% accuracy, 88.9 % +/- 12.1% precision, 80.0% +/- 14.4% recall (sensitivity), and 90.0% +/- 10.9% specificity. The mean AUROC was 85.0% +/- 9.0% and the mean AUPRC was 89.4% +/- 7.9%.

With dense optical flow, the mean performance metrics were 85.0% +/- 9.2% accuracy, 81.8% +/- 13.1% precision, 90.0% +/- 10.9% recall (sensitivity), and 80.0% +/- 14.4% specificity. The mean AUROC was 85.0% +/- 9.0% and the mean AUPRC was 88.4% +/- 7.9%.

With Gaussian blurring, the mean performance metrics were 83.3% +/- 9.2% accuracy, 83.3% +/- 13.7% precision, 83.3% +/- 13.7% recall (sensitivity), and 83.3% +/- 13.7% specificity. The mean AUROC was 83.3% +/- 9.3% and the mean AUPRC was 87.5% +/- 8.5%.

### Item-level analysis

Table 1 displays the mean absolute deviation of the mean answer for each privacy condition from the mean answer for the baseline condition. This difference provides a measure of the privacy condition’s effect on annotation.

**Table 1.** The mean absolute deviation for each privacy condition from the baseline condition answers for the behaviors used in the autism classifiers. This difference provides a measure of the privacy condition’s effect on annotation quality.

|  | Mean Deviation<br>for Pixelation | Mean Deviation<br>for Dense<br>Optical Flow | Mean Deviation<br>for Gaussian<br>Blurring |
| --- | --- | --- | --- |
| Abnormal<br>Speech | 0.28 | 0.32 | 0.29 |
| Echolalia | 0.39 | 0.45 | 0.47 |
| Repetitive or<br>Odd Language | 0.25 | 0.30 | 0.26 |
| Expressive<br>Language and<br>Conversation | 0.29 | 0.41 | 0.33 |
| Eye Contact | 0.29 | 0.37 | 0.33 |
| Facial<br>Expressiveness | 0.25 | 0.32 | 0.29 |
| Social<br>Interaction<br>Initiation | 0.26 | 0.30 | 0.31 |
| Shares<br>Excitement | 0.34 | 0.32 | 0.37 |
| Aggressive<br>Behavior | 0.09 | 0.12 | 0.12 |

We found that pixelation resulted in smaller deviations from the unmodified video condition compared to dense optical flow and Gaussian blurring. In all but one behavioral annotation (for sharing excitement), the mean deviation for pixelation was less than for the other two privacy conditions. Dense optical flow, which provides maximal privacy, did not have a discernable difference from Gaussian blurring (private but less so), providing support for the use of dense optical flow in translational settings.

The annotation with the lowest deviation across all conditions was for displaying aggressive behavior. This makes intuitive sense and serves as a sanity check, as no aggressive behavior was displayed in any of the videos we presented.

None of the 9 behaviors used for the classifiers contained a mean deviation above 0.5. This means that the mean deviation is less than one half of the distance between one categorical ordinal variable representing symptom severity and the variable indicating one severity level higher (all questions contained 4 multiple choice options). We note that while these deviations are consistently small, the aggregation of these deviations results in more misclassifications (see *Results: Effect of privacy conditions*).

## Discussion and Conclusion

We explored the potential for global image transformations to provide privacy for video subjects while still preserving behavioral annotation quality. While no individual question was drastically degraded, some behavioral annotations were degraded more than others. Pixelation consistently resulted in less drastic degradations than blurring and optical flow. We also found that the classifier’s predictions from the crowd’s annotation of the unaltered videos were strongly correlated with clinician global impressions. A slightly less strong correlation persisted even after all privacy modifications we tested, providing evidence that the classifier’s output can potentially be considered as an estimation of clinical global autism impression scores even when annotations are provided for a privacy-preserved video stream.

There are several limitations to the current study. While the configuration of questions we asked workers and clinicians resulted in worse performance by the clinicians’ annotations, this could have been due to over-sensitivity by the classifier rather than anything the clinicians did wrong. We therefore do not make any claims about the performance of clinicians as compared to crowd workers. An interesting limitation is that the definition of autism tends to shift over time with evolving DSM criteria and clinical practices [32]. Because clinicians were providing annotations several years after the videos of children were recorded, it is possible that the children who did not qualify for a diagnosis at the time the videos were recorded would qualify for a diagnosis by the time the clinicians reviewed the videos.

There are several interesting avenues of future work. The annotations provided by crowd workers can potentially be used to train computer vision classifiers detecting behaviors relevant to autism detection such as emotion evocation [15, 19, 44], hand or head stimming [46], and abnormal eye contact. While humans are worse at detecting certain behavioral patterns from videos when privacy mechanisms are applied, it is possible that convolutional neural networks can more easily detect these features by learning subtle and nonlinear feature maps beyond human comprehension. As machine learning algorithms improve and relevant databases become more plentiful, the possibility of removing humans from the remote detection pipeline seems increasingly inevitable.

Future work should ensure that all methods work for all stakeholders and such methods should therefore be evaluated across races and ethnicities to ensure fair and unbiased A.I. While some machine learning methods may help account for biased datasets, no technique matches the benefit of using balanced data. Fair and balanced healthcare A.I. initiatives must explicitly recruit participants in equal numbers across all demographics served to allow for equitable services for all.

## Data Availability

Data can be provided at the request to the authors.

## Acknowledgements

We thank all crowd workers who participated in the study. We also thank the clinicians who provided annotations for all study videos.

These studies were supported by awards to DPW by the National Institutes of Health (1R21HD091500-01 and 1R01EB025025-01). Additionally, we acknowledge the support of grants to DPW from The Hartwell Foundation, the David and Lucile Packard Foundation Special Projects Grant, Beckman Center for Molecular and Genetic Medicine, Coulter Endowment Translational Research Grant, Berry Fellowship, Spectrum Pilot Program, Stanford’s Precision Health and Integrated Diagnostics Center (PHIND), Wu Tsai Neurosciences Institute Neuroscience: Translate Program, Spark Program in Translational Research, Stanford’s Institute of Human Centered Artificial Intelligence, the Weston Havens Foundation, as well as philanthropic support from Peter Sullivan. PW would like to acknowledge support from the Stanford Interdisciplinary Graduate Fellowship (SIGF).

